# Mass screening of asymptomatic persons for SARS-CoV-2 using saliva

**DOI:** 10.1101/2020.08.13.20174078

**Authors:** Isao Yokota, Peter Y Shane, Kazufumi Okada, Yoko Unoki, Yichi Yang, Tasuku Inao, Kentaro Sakamaki, Sumio Iwasaki, Kasumi Hayasaka, Junichi Sugita, Mutsumi Nishida, Shinichi Fujisawa, Takanori Teshima

## Abstract

**Background:** COVID-19 has rapidly evolved to become a global pandemic due largely to the transmission of its causative virus through asymptomatic carriers. Detection of SARS-CoV-2 in asymptomatic people is an urgent priority for the prevention and containment of disease outbreaks in communities. However, few data are available in asymptomatic persons regarding the accuracy of PCR testing. Additionally, although self-collected saliva has significant logistical advantages in mass screening, its utility as an alternative specimen in asymptomatic persons is yet to be determined.

**Methods:** We conducted a mass-screening study to compare the utility of nucleic acid amplification, such as reverse transcriptase polymerase chain reaction (RT-PCR) testing, using NPS and saliva samples from each individual in two cohorts of asymptomatic persons: the contact tracing cohort and the airport quarantine cohort.

**Results:** In this mass-screening study including 1,924 individuals, the sensitivity of nucleic acid amplification testing with nasopharyngeal and saliva specimens were 86% (90%CI:77-93%) and 92% (90%CI:83-97%), respectively, with specificities greater than 99.9%. The true concordance probability between the nasopharyngeal and saliva tests was estimated at 0.998 (90%CI:0.996-0.999) on the estimated airport prevalence, 0.3%. In positive individuals, viral load was highly correlated between NPS and saliva.

**Conclusion:** Both nasopharyngeal and saliva specimens had high sensitivity and specificity. Self-collected saliva is a valuable specimen to detect SARS-CoV-2 in mass screening of asymptomatic persons.

## Introduction

Since its discovery in Wuhan, China in late 2019, the severe acute respiratory syndrome coronavirus 2 (SARS-CoV-2) has rapidly created a global pandemic of coronavirus disease 2019 (COVID-19). The fast evolution of this pandemic has been attributed to the majority of transmissions occurring through people who are presymptomatic or asymptomatic[1-3]. Accordingly, detection of the virus in asymptomatic people is a problem that requires urgent attention for the prevention and containment of the outbreak of COVID-19 in communities[4]. Currently, the diagnosis of COVID-19 is made by the detection of the nucleic acids of SARS-CoV-2 typically by real-time quantitative reverse transcriptase polymerase chain reaction (qRT-PCR) testing of specimens collected by nasopharyngeal swabs (NPS)[5, 6]. However, few data are available regarding the accuracy of qRT-PCR testing in asymptomatic persons upon which the implications of the current testing strategy depend. The sensitivity and specificity of PCR testing need to be elucidated in order to save unnecessary quarantine and contact-tracing, while minimizing new infections from presymptomatic persons.

Recently, specimen collection by NPS has been under scrutiny, as this method requires specialized health care workers and the use of personal protective equipment (PPE) to mitigate the risk of viral exposure. Consequently, self-collected saliva has been reported to have several advantages over NPS. As the name implies, self-collection of saliva eliminates the close contact in sampling, obviating the need for PPE. Additionally, providing saliva is painless and minimizes discomfort for the test subject. However, although we and others have shown the value of saliva as a diagnostic specimen in symptomatic patients[7-12], the utility of saliva in detecting the virus in asymptomatic persons remains to be elucidated.

## Methods

We conducted a mass-screening study to determine and compare the sensitivity and specificity of nucleic acid amplification using paired samples (self-collected saliva and NPS) for the detection of SARS-CoV-2 in two cohorts of asymptomatic individuals.

### Design and Population

The contact-tracing (CT) cohort included asymptomatic persons that have been in close contact with clinically confirmed COVID-19 patients with a positive qRT-PCR by NPS. Subjects in the CT cohort participated between June 12 and July 7, 2020 at several centres in Japan. Asymptomatic travelers arriving at Tokyo and Kansai international airports were enrolled from June 12 to June 23, 2020 as a separate cohort (airport quarantine (AQ) cohort). In both cohorts, all subjects were requested to provide NPS and saliva samples. All NPS samples in the CT cohort were tested by qRT-PCR. The NPS samples in the AQ cohort was tested by either qRT-PCR or reverse transcriptase loop-mediated isothermal amplification (RT-LAMP)[13, 14] at the discretion of the airport quarantine. All saliva samples in both cohorts were subjected to both qRT-PCR and RT-LAMP testing. This study was approved by the Institutional Ethics Board (Hokkaido University Hospital Division of Clinical Research Administration Number: 020-0116) and informed consent was obtained from all individuals.

### Diagnostic test

Saliva was diluted 4-fold with phosphate buffered saline (PBS) and centrifuged at 2000 × g for 5 min to remove cells and debris. RNA was extracted from 200 ^L of the supernatant or nasopharyngeal swab samples using QIAsymphony DSP Virus/Pathogen kit and QIAamp Viral RNA Mini Kit (QIAGEN, Hilden, Germany). Nucleic acids of SARS-CoV-2 were detected by qRT-PCR or RT-LAMP. qRT-PCR tests were performed, according to the manual by National Institute of Infectious Diseases (NIID, https://www.niid.go.jp/niid/images/epi/corona/2019-nCoVmanual20200217-en.pdf). Briefly, 5uL of the extracted RNA was used as a template. One step qRT-PCR was performed using THUNDERBIRD® Probe One-step qRT-PCR Kit (TOYOBO, Osaka, Japan) and 7500 Real-time PCR Systems (Thermo Fisher Scientific, Waltham, USA). The cycle threshold (Ct)-values were obtained using N2 primers (NIID_2019-nCOV_N_F2, NIID_2019-nCOV_N_R2) and a probe (NIID_2019-nCOV_N_P2). RT-LAMP was carried out to detect SARS-CoV-2 RNA using Loopamp®J 2019-SARS-CoV-2 Detection Reagent Kit (Eiken Chemical, Tokyo, Japan). The final reaction volume containing 10^l of viral RNA extract and 15^l of Primer Mix containing SARS-CoV-2 specific primers was dispensed into a reaction tube with dried amplification reagents including Bst DNA polymerase and AMV reverse transcriptase. This tube was incubated at 62.5°C with turbidity readings (optical density at 650 nm) and monitored for 35 minutes using the Loopamp Real-time Turbidimeter (Eiken Chemical Co., Ltd.,).

### Statistical analysis

Test value of qRT-PCR and RT-LAMP methods were illustrated by scatter plots and Kendall's coefficient of concordance *W* as nonparametric intraclass correlation coefficient taken non-linearity and censored value into consideration. The performance of diagnostic tests was evaluated by sensitivity *Se*_NPS_ (NPS)/ *Se*_saliva_ (saliva) and specificity *Sp*_NPS_ (NPS)/ *Sp*_saliva_ (saliva). Sensitivity was positive probability in infected population and specificity was negative probability in non-infected population. To evaluate the concordance between NPS and saliva test, true concordance probability was defined by *p* × *Se_NPS_* × *Se_saliva_*, × (1 − *p*) × *Sp_NPS_* × *Sp_saliva_*, that *p* was the prevalence of SARS-CoV-2.

The *Se*_NPS_, *Se*_saliva_, *Sp*_NPS_, *Sp*_saliva_ and *p* were jointly estimated using a Bayesian latent class model[15-17] since this method accounts for change of plans, rare positive cases. The prior distribution of specificity *Sp*_NPS_, *Sp*_saliva_ were *Beta*(201,1), reflecting the results of the in-hospital screening, all negative in more than 200 consecutive individuals with none subsequently developing COVID-19 (data not shown). The prior distribution of *Se*_NPS_, *Se*_saliva_ and *p* were *Beta*(1,1). The corresponding true concordance probability was estimated under varying prevalence values. For a sensitivity analysis, we estimated the true concordance probability when we imposed the constraint that the sensitivity of saliva test was equal to and 10% less than the sensitivity of NPS test.

Sample size in the CT cohort was calculated as 250 based on the prevalence of 0.1 and 25 positive samples were needed in order to keep the width of the 90% credible interval of sensitivity within 0.3 under the sensitivity at 0.7. Sample size in the AQ cohort was calculated 1,818 based on the probability that 90% credible interval of specificity over 99.0% would be 0.8 (likes statistical power) under the expected specificity being 99.5%.

The point estimate and 90% credible interval were used for the median and 5th to 95th percentile, respectively. All statistical analyses were conducted by SAS® Ver 9.4(Cary, NC).

## Results

### Demographics

Of the 2,558 persons screened, consent was obtained from 2,035 persons (80%) and 1,924 persons were included for analysis (Figure 1). The most common reason for exclusion was the presence of symptoms (n=95; 33%) and declined consent (n=493; 22%) in the CT and AQ cohorts, respectively. Only 16 persons (0.78%) were excluded due to insufficient saliva volume, confirming the feasibility of self-collection. Background characteristics of the 161 and 1,763 persons in the CT and AQ cohorts, respectively, are shown in Table 1. In the CT cohort, age and gender data were not made available from many subjects due to procedural reasons. This population mainly consisted of relatively young people between 20 and 50 years old. In the AQ cohort, the number of participants by the last point of embarkation was 467 (26%) from Europe (Amsterdam, Frankfurt, and London), 583 (33%) from Asia and Oceania (Bangkok, Jakarta, Manila, Seoul, Shanghai, Sydney, and Taipei), and 713 (40%) from North America (Chicago, Los Angeles, Seattle, and Vancouver). Because of the reduced number of international flights during this period, passengers from Central and South Americas, Africa, and the Middle East may have arrived via transit through any of the aforementioned regions.

**Figure 1.**
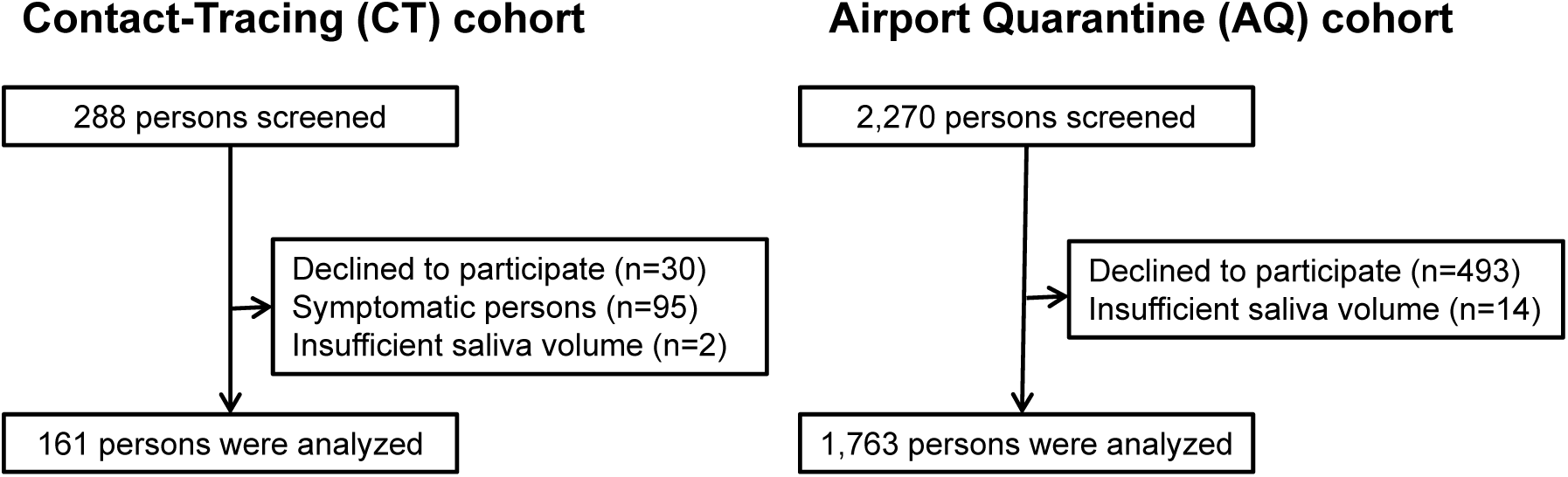
Flow diagram of participants

**Table 1.** Background characteristics

|  |  | contact-tracing cohort | airport cohort |
| --- | --- | --- | --- |
|  |  | N (%) | N (%) |
| Sex |  |  |  |
|  | Female | 26 (16.1) | 832 (47.2) |
|  | Male | 44 (27.3) | 927 (52.6) |
|  | unknown | 91 (56.5) | 4 (0.2) |
| Age |  |  |  |
|  | Median [IQR] | 44.9 [29.8, 66.4] | 33.5 [22.6, 47.4] |
|  | -19 | 2 (1.2) | 299 (17.0) |
|  | 20-29 | 16 (9.9) | 433 (24.6) |
|  | 30-39 | 13 (8.1) | 344 (19.5) |
|  | 40-49 | 9 (5.6) | 324 (18.4) |
|  | 50-59 | 8 (5.0) | 230 (13.0) |
|  | 60-69 | 9 (5.6) | 97 (5.5) |
|  | 70- | 13 (8.1) | 34 (1.9) |
|  | unknown | 91 (56.5) | 2 (0.1) |
| Last point of embarkation |  |  |  |
|  | North America | - | 713 (40.4) |
|  | Asia and Oceania | - | 583 (33.1) |
|  | Europe | - | 467 (26.5) |

### Sensitivity, Specificity and True concordance

In the CT cohort, SARS-CoV-2 was detected in 41 NPS samples and in 44 saliva samples, of which 38 individuals had both samples test positive (Table 2a). 114 persons were negative in both tests, which resulted in 152 of 161 matches. In the AQ cohort, viral RNA was detected in NPS and saliva in five and four samples, respectively, out of 1763 individuals (Table 2b).

**Table 2.** Diagnostic results of nasopharyngeal swab (NPS) and saliva test

**(a) Contact-tracing cohort (n=161)**
| NPS | saliva |  |
| --- | --- | --- |
|  | positive | negative |
| positive | 38 | 3 |
| negative | 6 | 114 |

**(b) Airport Quarantine cohort (n=1,763)**
| NPS | saliva |  |
| --- | --- | --- |
|  | positive | negative |
| positive | 4 | 1 |
| negative | 0 | 1758 |

The sensitivity of NPS and saliva were 86% (90% CI: 77-93%) and 92% (90% CI: 83-97%), respectively (Figure 2a), and the specificity of NPS and saliva were 99.93% (90% CI: 99.77-99.99%) and 99.96% (90%CI: 99.85-100.00%), respectively (Figure 2b). The estimated prevalence at the CT and AQ cohort was 29.6% (90%CI: 23.8-35.8%) and 0.3% (90%CI: 0.1-0.6%), respectively. The true concordance probability was estimated at 0.998 (90% CI: 0.996-0.999) in the AQ cohort. As shown in Figure 3, when the prevalence was varied from 0% to 30%, the point estimate for the true concordance probability ranged from 0.934 to 0.999 and the lower limit of the 90% CI was never below 0.9. True concordance probability with varying estimation constraints of sensitivity is shown to be very high (supplement 1), and therefore the qRT-PCR results from saliva and NPS appeared to be sufficiently consistent.

**Figure 2.**
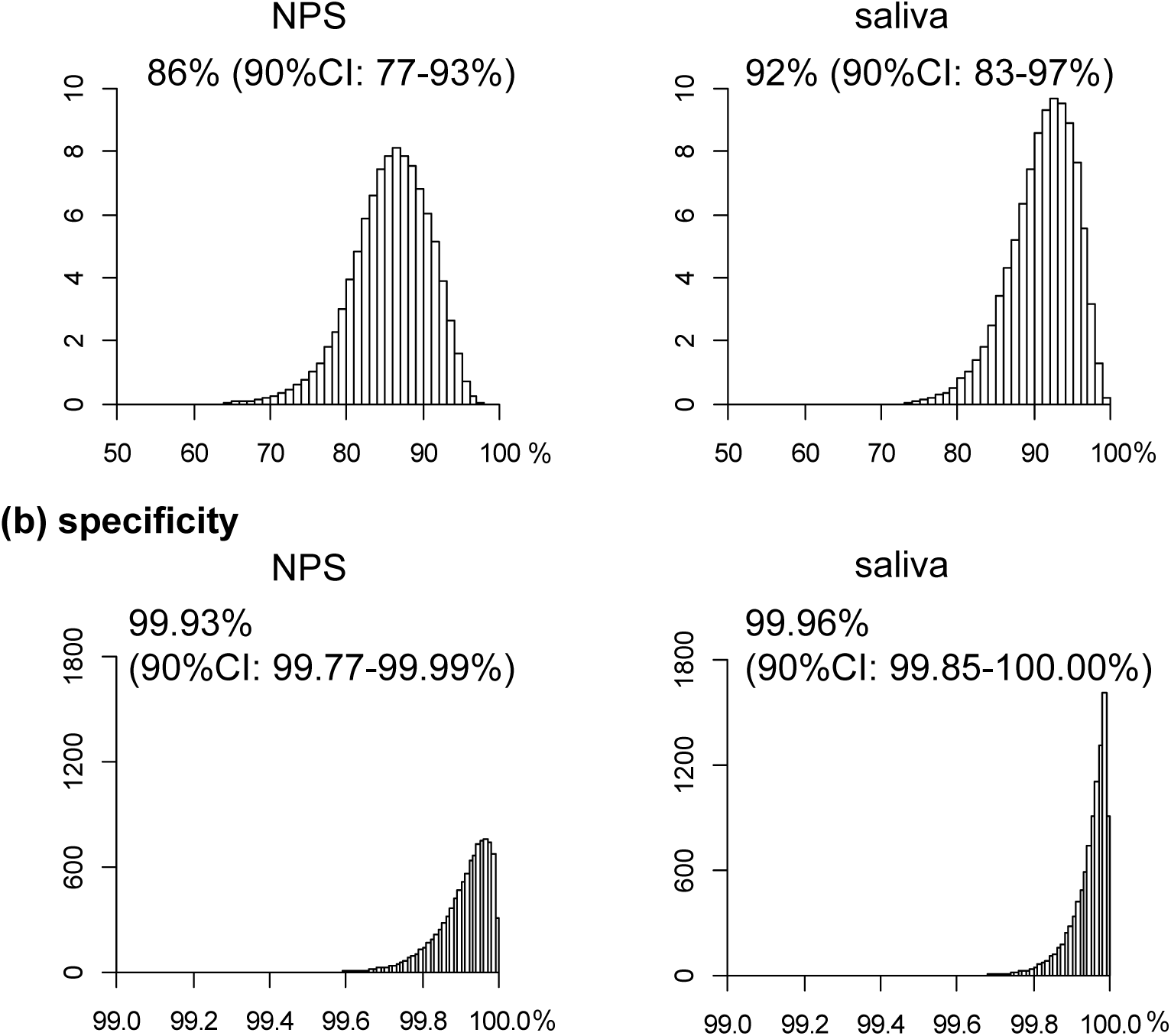
The sensitivity and specificity of nasopharyngeal swab and saliva. Histograms of posterior distribution of (a) sensitivity and (b) specificity. Point estimates and 90% credible interval (90%CI) defined by 5th to 95th percentile are shown.

**Figure 3.**
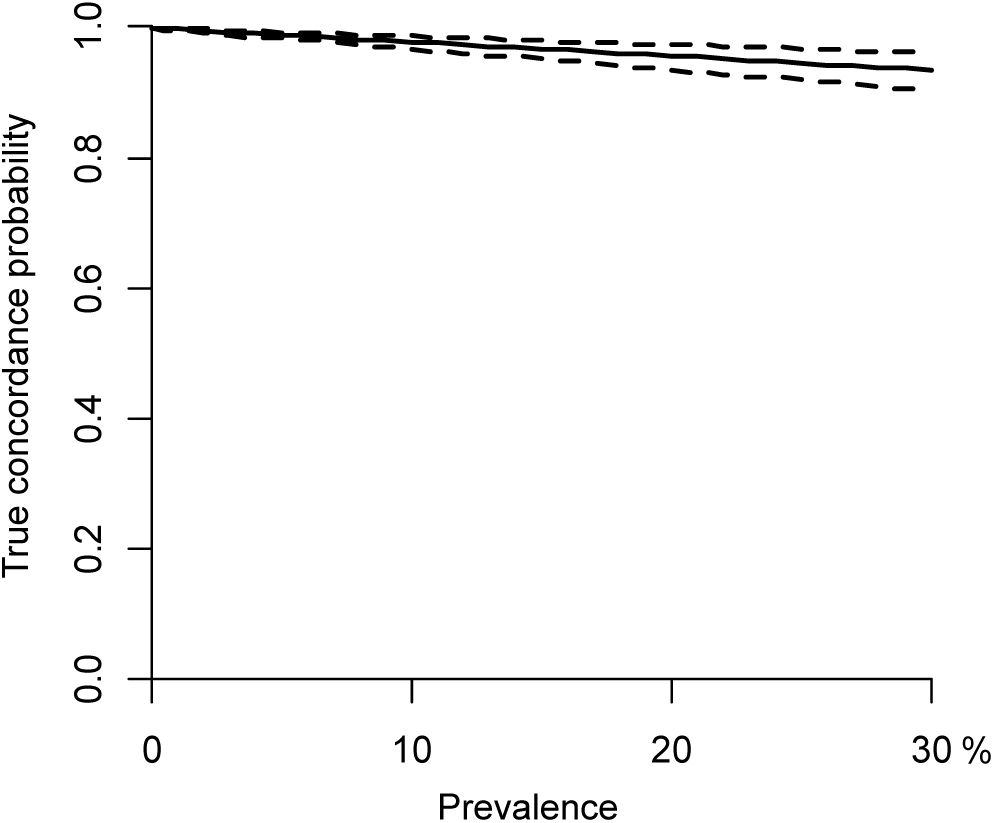
True concordance probability with varying rates of prevalence. The true concordance probability of diagnosis between nasopharyngeal swab and saliva test in populations with various prevalence. Solid line indicates point estimates and dashed lines indicate 90% credible interval.

### Comparison of the viral load between NPS and saliva samples

Scatter plot of the C_t_ values of qRT-PCR from the 45 positive specimens (either NPS or saliva) is depicted in Figure 4a. All three samples that were negative by saliva and positive by NPS had C_t_ values of 40 on NPS qRT-PCR test. On the other hand, six samples that were negative by NPS and positive by saliva had C_t_ values between 33.7 and 37.2 by saliva qRT-PCR. Kendall’s coefficient of concordance was 0.87, indicating that the viral load was equivalent between NPS and saliva samples.

**Figure 4.**
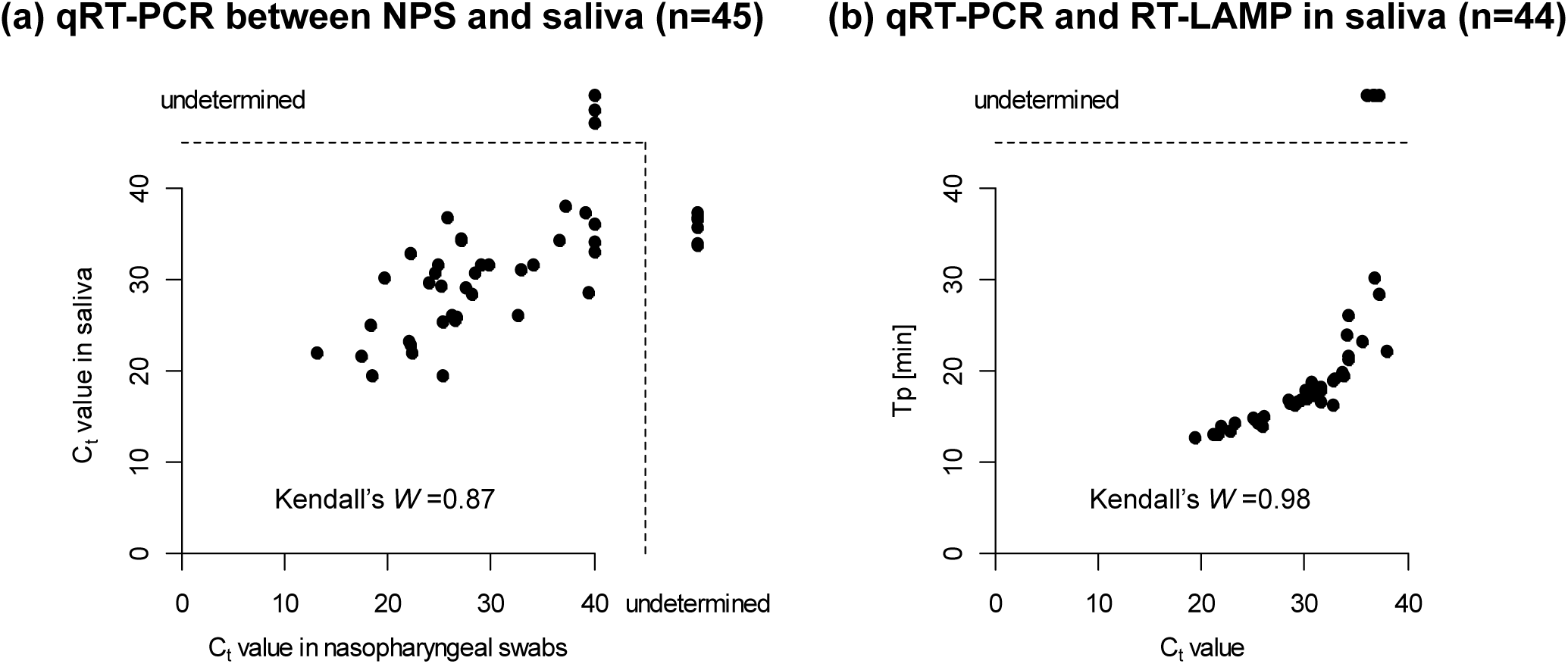
Comparison of the viral load between NPS and saliva. (a) C_t_ values determined with the qRT-PCR test of nasopharyngeal swab and saliva are plotted. (b) Times to detecting positive results (Tp) determined by the RT-LAMP test of saliva are plotted against C_t_ values determined by the qRT-PCR test of saliva. *W* indicates Kendall’s coefficient of concordance. Data were plotted with one of the tests being positive and the values being measured.

### Test values of RT-PCR and RT-LAMP methods

To confirm the equivalence of the qRT-PCR and RT-LAMP methods, a scatter plot of time for detecting positive results (Tp) with RT-LAMP against C_t_ values of qRT-PCR test using 44 saliva samples is shown in Figure 4b. Four samples that were negative by RT-LAMP and positive by qRT-PCR had C_t_ values ranging from 36.0 to 37.3, indicating very low viral loads (Kendall's coefficient of concordance = 0.98). Excluding these four samples, concordance between qRT-PCR and RT-LAMP was demonstrated in saliva specimens in 87 samples (36 positive and 51 negative) in the CT cohort. In the AQ cohort, all 1763 samples (4 positive and 1759 negative) were concordant.

## Discussion

This study examined the accuracy of detecting SARS-CoV-2 by qRT-PCR using NPS and saliva in a significant number (n=1,924) of asymptomatic individuals. Our results showed that qRT-PCR in both specimens had specificity greater than 99.9% and sensitivity approximately 90%, validating the current practice of detecting infection by nucleic acid amplification.

We report for the first time the accuracy of viral detection using natural clinical specimens of asymptomatic persons[18], that the sensitivity is higher than the 52% to 71% reported in symptomatic patients[5, 19-22]. COVID-19 literature to date have been consistent in identifying the peak viral load at symptom onset with subsequent decline[7, 19, 23-26], suggesting the possibility of higher presymptomatic viral load. More recent studies have also shown that infectiousness peaks on or before symptom onset[27], and that live virus can be isolated from asymptomatic individuals[28]. Concomitantly, there have been reports of discrepancy between viral load as detected by qRT-PCR and contagiousness[28-30], which may be of utmost importance in controlling outbreaks, as the potential to infect close contacts lends credibility to the current strategy of self-quarantine. Although the relationship of contagiousness and viral load is a subject in need of further investigation, abrogation of early infectiousness may also be an effective drug development target.

The current study further extends that saliva may be a beneficial alternative to nasopharyngeal fluid in detecting SARS-CoV-2 in asymptomatic carriers. The comparison between paired samples have shown equivalent utility with similar sensitivity and specificity. However, self-collected saliva has significant advantages over NPS sampling especially in the setting of mass screening. For example, saliva collection is non-invasive and does not require specialized personnel nor the use of PPE, which saves time and cost. Additionally, providing saliva is painless and minimizes discomfort for the patient. These significant advantages became immediately apparent during our sample collection at the airport quarantine, where queue of international arrivals filtered smoothly through multiple collection booths. Obtaining saliva is simply more conducive to simultaneous mass screening of large number of individuals, in settings such as social and sporting events.

Previous studies comparing the viral load between NPS and saliva samples report conflicting results. Wyllie et al. showed that the viral load was five-times higher in saliva than NPS[23], while others have reported results to the contrary[9, 26]. Our results clearly show the viral loads to be equivalent between NPS and saliva in asymptomatic individuals and both specimens may be useful in detecting viral RNA.

Among the limitations of any diagnostic modality is the possibility of obtaining false results with serious consequences. While persons infected with SARS-CoV-2 with falsely negative test may be left in society without the necessary precautions to keep him/her from transmitting the virus, false positive non-infected persons may undergo unnecessary quarantine and labour-intensive contact tracing measures. Although the high specificity of qRT-PCR reported herein may be reassuring in individual cases, the implications of mass testing depends on the prevalence of disease in the subject population. However, point prevalence is unknowable *a priori* and extremely difficult to assess in rapidly evolving outbreaks from carriers with relatively long presymptomatic periods. Rather, insights on mass testing may be gained through carefully monitoring test positivity in relation to the total number of tests performed. For example, with greater than 99.9% specificity, a positive result in five percent of all tests would indicate that more than 4.9% (out of the 5%) are true positives, with a positive predictive value (PPV) of at least 98%. On the other hand, if only 0.3% of all tests return positive (e.g. in isolated localities with very few disease), the PPV would be (0.3%-0.1%)/0.3% = 0.67, erroneously labelling one third of all positive tests. As PPV is dependent on the prevalence of disease, mass testing using a highly specific test will remain effective as long as test positivity remains relatively high.

RT-LAMP is an isothermal nucleic acid amplification technique that allows results to be obtained in approximately 30-60 minutes and a recent study showed the equivalent efficacy of RT-PCR and RT-LAMP in symptomatic patients [12]. In this study, we confirmed this in a large population of asymptomatic persons using saliva samples; there were no samples that were negative by NPS RT-LAMP and positive by saliva. It is unlikely that the sensitivity of the RT-LAMP method is significantly less than that of qRT-PCR, and the RT-LAMP testing has little impact on our conclusions. Our study suggests that RT-LAMP is a useful alternative to RT-PCR for the diagnosis of SARS-CoV-2.

The current study lacks longitudinal data and clinical confirmation of positive cases. Nonetheless, this is the first study in asymptomatic individuals comparing paired samples of NPS and saliva. Rapid detection of asymptomatic infected patients is critical for the prevention of outbreaks of COVID-19 in communities and hospitals. Mass screening of the virus using self-collected saliva can be performed easily, non-invasively, and with minimal risk of viral transmission to health care workers.

## Data Availability

The datasets generated during and/or analysed during the current study are not publicly available due to unaccepted work on any peer-reviewed journal but will be published after acceptance of peer-reviewed journal.

## Contributors

IY, KS, JS, MN and TT determined the study design. IY, PS, YU, SI, KH, MN, SF and TT collected the data. IY, KO, YU, YY, TI, KS did statistical analysis. IY, PS, TT drafted the manuscript and all authors reviewed critically and approved the final manuscript.

## Acknowledgement

We thank Tokyo airport quarantine station and Kansai airport quarantine station for cooperation; Megumi Aoki, Miwa Aoki, Nana Arai, Satomi Araki, Cao Cuicui, Kazumi Hasegawa, Masato Horiuchi, Dr. Nao Kurita, Dr. Aki Nakamura, Chiho Okabe, Mana Okamura, Yusuke Sakai, Dr. Akahito Sako, Natsumi Satake, Maki Shimatani, Kaki Tanaka, Maina Toguri, Sachiko Tominaga and Hana Wakasa for assistance in collecting saliva samples.

**Supplement 1**. True concordance probability under several scenarios.

